# Estimating the undetected infections in the Covid-19 outbreak by harnessing capture-recapture methods

**DOI:** 10.1101/2020.04.20.20072629

**Authors:** Dankmar Böhning, Irene Rocchetti, Antonello Maruotti, Heinz Holling

## Abstract

A major open question, affecting the policy makers decisions, is the estimation of the *true* size of COVID-19 infections. Most of them are undetected, because of a large number of asymptomatic cases. We provide an efficient, easy to compute and robust lower bound estimator for the number of undetected cases. A “modified” version of the Chao estimator is proposed, based on the cumulative time-series distribution of cases and deaths. Heterogeneity has been accounted for by assuming a geometrical distribution underlying the data generation process. An (approximated) analytical variance formula has been properly derived to compute reliable confidence intervals at 95%. An application to Austrian situation is provided and results from other European Countries are mentioned in the discussion.

## 1 Introduction

Currently, health systems across the globe are challenged by the ongoing Covid-19 pandemic. It is not a simple task to assess the efficiency of current health systems in detecting, treating, and preventing onward transmission of Covid-19, as the number of undetected infections is by definition unknown. Understanding the diffusion and assessing the number of real infections of Covid-19 is crucial for implementing effective public and health policies in tackling the virus. Unfortunately, official reporting and statistics significantly underestimate the *true* number since there exists a vast proportion of asymptomatic infected patients including those with mild symptoms among all infected individuals who are not detected. Indeed, the infected individuals with low-mild symptoms are likely not going to get in contact with the health care system and will also not be recorded in official statistics.

For example, reports estimate the number of infected in Italy to be around 3.5 times higher than reported [Tuite et al.(2020)]. Slightly lower estimates have been given for Germany [Ranjan(2020)]. Another study discusses that Italy mostly focuses on testing in hospitals with symptoms; hence, the roughly 50% asymptomatic are not covered by this approach [Onder et al.(2020)]. The same percentage of asymptomatic is also reported in Iceland [Shahan(2020)]. The asymptomatic individuals in fact can be a direct transmitter of the virus and their unawareness can indirectly strengthen and increase the transmission of Covid-19. Indeed, it seems fair to say that the undetected cases are the major dirver in spreading the disease as detected cases are and will be systematically contained.

Most of the existing analyses performed a secondary data analysis from several sources of data already in the public domain [Menkir et al.(2020)]. Because published estimates of the distribution of Covid-19 vary widely, with estimates of the basic reproduction number, R0, alone ranging from subcritical (i.e., 1) to 3 [Li et al.(2020), Zhao et al.(2020), Zhou et al.(2020)], mathematical models of infectious diseases, such as Susceptible-Infected-Recovered models, computing the theoretical number of people infected with a contagious illness in a closed population over time, needs to be evaluated on a range/grid of simulated values, each based on different assumptions and adjusted based on data from different geographic areas [Chen et al.(2020)]. Other much simpler [Nishiura et al.(2020)] or sophisticated [Flaxman et al.(2020)] approaches are also used to estimate the number of undetected cases, but with large, almost unacceptable, uncertainty on the obtained estimates.

The purpose of this contribution is to propose a lower bound estimator for the number of people affected by Covid-19 but not detected for various reasons, the major one being that they are asymptomatic. In other words, the aim is to estimate the size of an elusive, i.e. partially unobserved, population. Capture-Recapture (CR) methods are designed to achieve this goal. Our proposal is developed using the cumulative distribution of the observed cases and deaths. The use of CR methods is not straightforward as we are dealing with an *open* population, subject to deaths, and heterogeneity in the probability of being detected. A data-modified version of Chao’s estimator under a geometric distribution is introduced. It accounts for heterogeneity in a simple way and can be easily computed starting from data collected by all government sources. In this way, the policy makers can have benchmark, statistically valid, estimates of the lower bound for the total number of cases and, accordingly, adjust their interventions.

This short note is organized as follows. In section 2 we introduce the basic notation and how we are going to work with the cumulative distribution of observed cases and deaths. Section 3 provides all the necessary details to obtain the estimates. An example to Austrian data is provided. A discussion showing other interesting insights concludes.

## 2 Basic notation

We will denote with *N*(*t*) the cumulative count of infections at day *t* where *t*_0_ = *t*_0_, …,*t*_*m*_. Hence Δ*N*(*t*) = *N*(*t*) − *N*(*t*−1) are the number of new infections at day *t* where *t* = *t*_0_ + 1, ⋯, *t*_*m*_. Also, let *D*(*t*) denote the cumulative count of deaths at day *t* where *t* = *t*_0_ + 1,…,*t*_*m*_. *t*_0_ defines the beginning of the observational period and *t*_*m*_ defines the end. We assume the trivial assumption *t*_*m*_ > *t*_0_, so that the observational window is not empty. Again, we denote with Δ*D*(*t*) = *D*(*t*) − *D*(*t*−1) the count of new deaths at day *t* where *t* = *t*_0_ + 1, …, *t*_*m*_. To illustrate, we look at these data (taken from https://www.worldometers.info/coronavirus/country/austria/) for the country of Austria as provided in Table 1 for the infections and in Table 2 for the deaths:

**Table 1:**
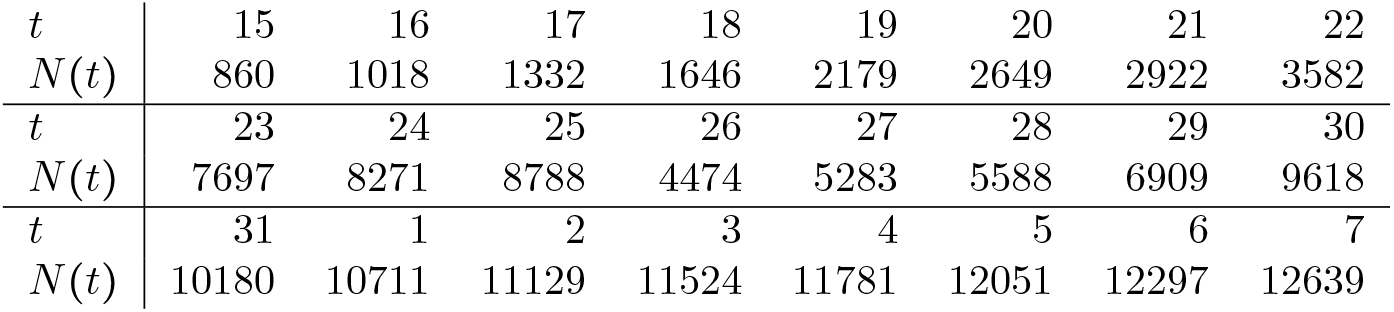
Cumulative counts of infections with Covid19 for Austria starting at *t*_0_ = 15 March 2020 to *t*_*m*_ = 7 April 2020

|  |  |  |  |  |  |  |  |  |
| --- | --- | --- | --- | --- | --- | --- | --- | --- |
| $t$ | 15 | 16 | 17 | 18 | 19 | 20 | 21 | 22 |
| $N(t)$ | 860 | 1018 | 1332 | 1646 | 2179 | 2649 | 2922 | 3582 |
| $t$ | 23 | 24 | 25 | 26 | 27 | 28 | 29 | 30 |
| $N(t)$ | 7697 | 8271 | 8788 | 4474 | 5283 | 5588 | 6909 | 9618 |
| $t$ | 31 | 1 | 2 | 3 | 4 | 5 | 6 | 7 |
| $N(t)$ | 10180 | 10711 | 11129 | 11524 | 11781 | 12051 | 12297 | 12639 |

**Table 2:** Cumulative counts of deaths from Covid19 for Austria starting at *t*_0_ = 15 March 2020 to *t*_*m*_ = 7 April 2020

|  |  |  |  |  |  |  |  |  |  |  |  |  |
| --- | --- | --- | --- | --- | --- | --- | --- | --- | --- | --- | --- | --- |
| $t$ | 15 | 16 | 17 | 18 | 19 | 20 | 21 | 22 | 23 | 24 | 25 | 26 |
| $D(t)$ | 1 | 2 | 4 | 4 | 6 | 6 | 8 | 16 | 21 | 28 | 31 | 49 |
| $t$ | 27 | 28 | 29 | 30 | 31 | 1 | 2 | 3 | 4 | 5 | 6 | 7 |
| $D(t)$ | 58 | 68 | 86 | 108 | 128 | 146 | 158 | 168 | 186 | 204 | 220 | 243 |

## 3 Linking with the capture-recapture methodology

The question arises how this can be linked to a capture-recapture approach. For this purpose we briefly review the capture-recapture model we like to harness here. Suppose a target population is sampled for units of interest repeatedly. Let *X* denote the number of times a unit is identified in this sampling process. Also, let *p*_*x*_ denote the probability of identifying a unit *x* times where *x =* 0…. In the capture-recapture world the following mixture model is quite common:

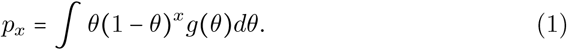

In (1) occurs the geometric distribution as a suitable count distribution and its parameter is allowed to experience population heterogeneity (as expressed by the density *f* (*θ*)) to reflect varying identification probabilities across the target population. Often the Poisson distribution is used in (1) instead of the geometric distribution. However, we prefer to use the latter as we think of the geometric distribution as a Poisson distribution mixed with an exponential density, hence able to incorporate already some of the likely present heterogeneity in the populaiton.

Using the Cauchy-Schwarz inequality for moments, it is possible to show that for the probability *p*_0_ of missing a unit of interest the following inequality holds:

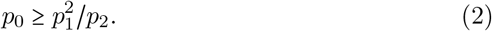

Replacing *p*_1_ and *p*_2_ on the right-hand side of (2) with the observed frequencies *f*_1_ of those identified exactly once and *f*_2_ of those identified exactly twice leads to the lower bound estimate of Chao [Chao(1987), Chao(1989), Chao and Colwell(2017)]:

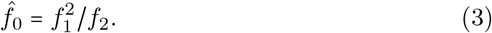

Here *f*_0_ is frequency of units that remain unobserved or hidden.

The idea is to apply this estimator (3) day-wise. We take an arbitrary day *t*. At this day we have Δ*N*(*t*) new infections. This will be viewed as *f*_1_, the infected people identified just once. If we look at Δ*N* (*t* − 1), then this is the count of new infections the day before. But these will still be infected at day *t* unless they decease. So, *f*_2_ corresponds to Δ*N* (*t* − 1) − Δ*D*(*t*). We can ignore the number of recoveries as we are looking at infections which are very recent (notified at day *t* or *t*−1). Hence we are able to give the estimate for the number of hidden infections at day *t* as

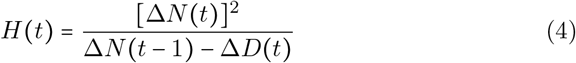

and global estimate of hidden infections is achieved by summing up over all days in the observational period:

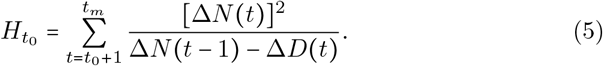

We will use a bias-corrected form of (4) suggested by [Chao(1989)] and given as

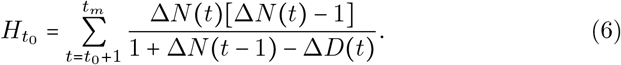

We define the understanding that Δ*N*(*t*−1)− Δ*D*(*t*) is set to 0 if it becomes negative, in other words we use *max* {0, Δ*N* (*t* −1) − Δ*D* (*t*)}. The final estimate of the total size of infection is then given as what has been observed at the end of the observational window *t*_*m*_ and the estimate of the hidden numbers:

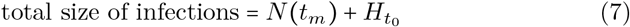

We need to address the uncertainty involved in the estimator (6). A variance estimate of (4) has been provided in [Niwitpong et al.(2013)] and is given here as

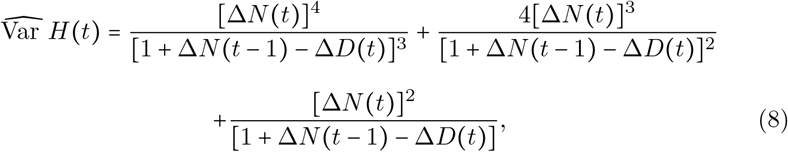

so that the final variance estimate of 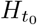 is given as

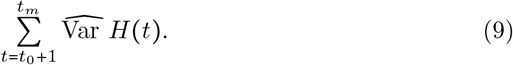

A 95% confidence interval can then be constructed by means of

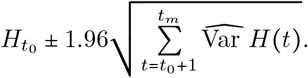

## 4 Application to the Austrian situation

We now apply (5) to the Austrian data. The results are provided in Table 3 which includes estimates of the hidden and total (observed + hidden) cases with 95% confidence intervals. At the 7th of April the number of infections was 12639 which is the observed number. We have chosen the 15th of March as beginning of the observational period. However other dates are possible as well so that we looked at estimates in dependence of the beginning of the observation period. It can be seen that results change slightly. Of course, if the window is made too small estimates of hidden numbers will only refer to observations made in this window. The major question arises if the estimates of Table 3 are realistic. For Austria we have an independent study on the size of the Covid19 outbreak (https://www.welt.de/politik/ausland/-article207187759/Coronavirus-Eisberg-hoeher-als-gedacht-Oesterreich-legt-Dunkelziffer-Studie-vor.html). The study was led by Christoph Fassmann and is known as the *dark number study*. The study was rolled out during the 1 April and 6 April 200 and sampled 1544 persons across Austria covering all ages up to 94 years. According to the study, the proportion of infected people was 0.0033. If this proportion is applied to the population of Austria, as study in media release points out, during the study period there were 28500 infected persons in Austria. The study estimates that we have provided matches very well with the results of the study, independent where we start the observational window. The dark number study also reports a 95% confidence interval for the proportion of infected persons which ranges from 0.0012 to 0.0076, corresponding to 10200 and 67400 infected persons, respectively. Clearly, the capture-recapture estimate is included in this large interval but as we are able to utilize much larger routinely collected data on infected persons the uncertainty provided by the capture-reacpture approach is considerably reduced which is reflected in the relative short confidence intervals.

**Table 3:** Estimated hidden and total cases of Covid19 for Austria starting and various sizes of the observational window ranging from *t*_0_ = 15 March 2020 to *t*_0_ = 18 March 2020; the second part of the table contains the associated proportions of total population in Austria (8.859 million)

| $t_0$ | hidden cases | total cases | 95% CI |
| --- | --- | --- | --- |
| 15 | 17264 | 29560 | 28412 – 30709 |
| 16 | 16638 | 28935 | 27800 – 30069 |
| 17 | 16326 | 28623 | 27491 – 29754 |
| 18 | 15420 | 27716 | 26602 – 28831 |
| 15 | 0.0019 | 0.0033 | 0.0032 – 0.0035 |
| 16 | 0.0019 | 0.0033 | 0.0031 – 0.0034 |
| 17 | 0.0018 | 0.0032 | 0.0031 – 0.0034 |
| 18 | 0.0017 | 0.0031 | 0.0030 – 0.0033 |

## 5 Discussion

The proposed method answers to a fundamental open question: “How many undetected cases are going around?”. Of course, we provide a lower bound, but this information may be treated as a starting point whenever interventions and tools to dampen the spread of the epidemic are rolled out. CR methods are easy to apply in practice, and this is one of the merits of the method. Moreover, we simply use time series of cumulated data, readily available from governments sources. Given that individual data are not publicly available, CR methods provide a straightforward solution to shed light on undetected cases, incorporating heterogeneity that may arise in the probability of being detected simply considering the widely known and used geometric distribution.

The example provided here relies on Austrian data, but many other Countries can be analyzed even if there are not benchmark survey studies to compare with. For example, taking data up to 18/04/2020 from https://github.com/open-covid-19/data on several European countries and considering data from the day which we record the first death, we obtain the estimates of undetected cases for Italy, Germany, Spain, UK and Greece (see Table 4). The last column in Table 4 shows the ratio of the total estimated cases to the observed cases. There is a remarkable stability around the value of 2.3.

**Table 4:** Estimated hidden and total cases of Covid-19 for several European countries, at 18/04/2020

| Country | hidden cases | total cases | 95% CI | total/observed |
| --- | --- | --- | --- | --- |
| Italy | 211768 | 384201 | 381649 – 386762 | 2.23 |
| Germany | 178451 | 315890 | 312429 – 319350 | 2.30 |
| Spain | 232057 | 423783 | 421112 – 426454 | 2.21 |
| UK | 149150 | 257842 | 255482–260202 | 2.37 |
| Greece | 2901 | 5108 | 4718–5499 | 2.31 |

All the obtained estimates are surrounded by some uncertainty. Confidence intervals for the “modified” Chao lower bound have been provided and are seemingly reliable, in particular compared to those presented in other studies. We emphasize that the estimates provided are conservative, in the sense that they provide lower bounds on the size of undetected infections. However, we have provided some evidence such as in the situation of Austria that these lower bound are not far away from the true size of infection in the target population. This needs to be followed up by further comparisons with representative sampling studies on target population infection.

This is just a first evidence on the use of capture-recapture methods to study Covid-19 data. Another question is still open: “is there a way of estimating an upper bound for the number of undetected cases?”. Again capture-recapture methods could be implemented to provide an answer to this question and help policy makers to evaluate the Covid-19 epidemic situation locally and at the current phase of its development.

## Data Availability

All data used are publically available and the sources have been identified in the paper.

## Notes

### Competing Interest Statement

The authors have declared no competing interest.

### Clinical Protocols

https://www.worldometers.info/coronavirus/

### Funding Statement

No funding received.

